# Clinical utility of plasma APOE proteotyping for Alzheimer’s disease risk and prognosis

**DOI:** 10.64898/2026.10.08.26364972

**Authors:** Ruyu Shi, Lamia T. Choity, Marissa F. Farinas, Xuemei Zeng, James E. Emanuel, Anum Saeed, Steven E. Reis, Oscar L. Lopez, Ilyas M. Kamboh, Ann D. Cohen, Ariel G. Gildengers, Kristine A. Wilckens, Thomas K. Karikari

## Abstract

**Background:** Alzheimer’s disease (AD) is a complex neurodegenerative disorder strongly influenced by genetic factors, with apolipoprotein E (APOE) ε4 allele being the most established risk variant. Although DNA-based genotyping remains standard, plasma proteotyping may provide scalable, isoform-specific alternatives. Here, we aimed to investigate the clinical utility of APOE proteotyping as a surrogate for genotyping in different clinical and research cohorts.

**Methods:** Plasma APOE4 protein were quantified using the NULISAseq CNS panel across four cohorts (ADRC, HeartSCORE, LATTICE, and ALPS). DNA-based APOE genotyping was the reference standard. Proteotyping thresholds for APOE ε4 carrier status and copy number were derived in ADRC and externally validated in the other cohorts. Logistic, proportional-odds, and Cox proportional-hazards models compared genotype- and proteotype-defined groups in relation to amyloid positivity, cross-sectional CDR global scores, and time to CDR-SB worsening, respectively.

**Results:** Plasma APOE4 accurately classified APOE ε4 carrier status in ADRC (AUC=0.977, 95%CI 0.967-0.985; threshold 5.78 NULISA protein quantification [NPQ] value) and achieved near-perfect external validation in HeartSCORE, LATTICE, and ALPS (all AUCs=1.000, 95%CI 1.000-1.000). Plasma APOE4 also well-differentiated heterozygous from homozygous ε4 carriers (ADRC AUC=0.879, 95%CI 0.834-0.921; threshold 12.23 NPQ value). Genotype- and proteotype-defined ε4 carrier status showed similar associations with amyloid positivity (OR=4.40 vs 4.45), higher CDR global scores (OR=1.50 for both), and CDR-SB worsening (HR=1.22 for both).

**Discussion:** Plasma APOE4 proteotyping accurately distinguished genotype-defined APOE ε4 carrier status and copy number while demonstrating AD-related associations comparable to DNA genotyping.

## 1 Background

Apolipoprotein E (APOE) is a lipid transport protein that plays a central role in cholesterol metabolism and neuronal maintenance.^1^ It exists as three common isoforms, ε2, ε3, and ε4, defined by amino acid substitutions at residues 112 and 158, and is a major genetic determinant of Alzheimer’s disease (AD), a multifactorial neurodegenerative disorder accounting for 60%–80% of dementia cases and arising from a complex interplay of genetic and environmental factors.^2^ The APOE ε4 allele confers a dose-dependent increase in disease risk. Its population frequency varies across ancestral groups, ranging from 13% to 23% in individuals of European ancestry, 26% to 30% in those of African ancestry, and approximately 7% in Asian populations.^3^

Individuals carrying one ε4 allele have an approximately two- to three-fold increased risk of AD, whereas homozygous carriers may have a 10- to 15-fold greater risk, with penetrance varying across racial and ethnic groups.^4,5^ Mechanistically, APOE ε4 has been linked to multiple features of AD pathophysiology, including impaired β-amyloid (Aβ) clearance, increased tau hyperphosphorylation and neurofibrillary tangle formation, and altered brain glucose metabolism and insulin signaling.^6^ Recent evidence further suggests that APOE ε4 homozygosity is associated with near-complete penetrance of cerebral Aβ deposition by age 65, underscoring its central role in amyloid pathology.^7^

Apolipoprotein E (APOE) genotype is typically determined using single-nucleotide polymorphism (SNP)-based genotyping or sequencing in specialized laboratories, with buffy coat being the standard specimen source.^8,9^ Although germline variants such as APOE can be detected in plasma cell-free DNA, its fragmented nature and low abundance limit robustness compared with genomic DNA derived from buffy coat.^10^ In this context, plasma-based APOE proteotyping provides a practical and scalable alternative to conventional genotyping by directly quantifying APOE isoforms. Unlike DNA-based methods that require cellular material, this approach can be readily applied to archived plasma samples. Notably, APOE proteotyping has recently been incorporated into the NULISAseq (NUcleic acid Linked Immuno Sandwich Assay sequencing) CNS panel, an ultrasensitive multiplex immunoassay that is gaining increasing attention in AD biomarker research. The NULISAseq CNS panel aims to simultaneously measure core AD biomarkers (amyloid β, neurofibrillary tangles, and neurodegeneration) alongside markers of co occurring processes, including inflammation, vascular dysfunction, synucleinopathy, proteostasis impairment, cytoskeletal alterations, and nucleic acid dysregulation, thereby enabling a more comprehensive characterization of AD pathophysiological heterogeneity.^11–13^

To date, evidence supporting APOE proteotyping particularly using NULISAseq CNS panel has been limited, with prior evaluations conducted in a single cohort of modest size (∼170 participants).^14^ The generalizability and robustness of these findings therefore remain uncertain. In this study, we leveraged multiple independent cohorts to provide a more rigorous assessment of concordance between APOE proteotype and genotype across diverse populations and study settings. Importantly, samples were analyzed across different assay runs, NULISAseq CNS120 versions, and reagent lots, enabling evaluation of consistency and robustness under varying technical conditions.

## 2 Methods

### 2.1 Participants and data selection

Data were drawn from four available cohorts, including the Alzheimer’s Disease Research Center from University of Pittsburgh (ADRC; n=1408), Heart Strategies Concentrating on Risk Evaluation (HeartSCORE; n=139), Lithium as a Treatment to Prevent Impairment of Cognition in Elders (LATTICE; n=77), and Alzheimer’s Pathways Sleep Study (ALPS; n=34). The ADRC is a longitudinal memory clinic cohort based at the University of Pittsburgh that enrolls older adults across the cognitive spectrum from western Pennsylvania.^15^ The HeartSCORE is a community-based longitudinal study conducted in Allegheny County, Pennsylvania, which recruited baseline middle-aged participants at baseline to examine the racial disparities in cardiovascular disease (CVD) risk factors.^16^ Our study is based on a subset of HeartSCORE participants who were recruited for a neuroimaging study during their late-life visit.^17^ The LATTICE is a randomized clinical controlled trial conducted at the University of Pittsburgh to evaluate the efficacy of lithium treatment in adults aged ≥60 years with amnestic mild cognitive impairment (aMCI).^18^ The ALPS study is an experimental, randomized controlled trial conducted in the University of Pittsburgh School of Medicine evaluating the effects of a 4-week behavioral time-in-bed restriction intervention on slow-wave sleep, cognitive performance, and Alzheimer’s disease pathophysiology in older adults aged 65-85 years.^19^ Plasma samples were collected and available in 34 participants from the ALPS study, which was ongoing at the time of the current analysis. All four studies were approved by the University of Pittsburgh Institutional Review Board, and all participants provided written informed consent. Detailed descriptions of all studies have been reported in prior publications as cited above.

Age, sex (biological), race, and education level were obtained by self-report at the baseline visit. Participant characteristics stratified according to their APOE ε*4* carrier status and complete *APOE* genotype groups were shown in **Table 1** and **Supplementary Table S1**. Due to the small sample size for other races, only participants self-identified as non-Hispanic White (NHW) or Black/African American (B/AA) were included in the statistical analysis.

**Table 1.** Participant characteristics and plasma APOE4 levels by APOE ε4 copy number.

| Variable | N | Overall | APOE ε4<br>non-Carrier | Heterozygotic | Homozygotic | p-value<sup>a</sup> |
| --- | --- | --- | --- | --- | --- | --- |
|  |  | N = 1,658 | N = 940 | N = 600 | N = 118 |  |
| **Cohort** | 1,658 |  |  |  |  | NA |
| ADRC |  | 1,408 (84.9%) | 763 (81.2%) | 533 (88.8%) | 112 (94.9%) |  |
| ALPS |  | 34 (2.1%) | 27 (2.9%) | 6 (1.0%) | 1 (0.8%) |  |
| HeartScore |  | 139 (8.4%) | 103 (11.0%) | 34 (5.7%) | 2 (1.7%) |  |
| LATTICE |  | 77 (4.6%) | 47 (5.0%) | 27 (4.5%) | 3 (2.5%) |  |
| **APOE Genotype** | 1,658 |  |  |  |  | NA |
| ε2/ε2 | | 8 (0.5%) | 8 (0.9%) | 0 | 0 | |
| ε2/ε3 | | 141 (8.5%) | 141 (15.0%) | 0 | 0 | |
| ε2/ε4 | | 56 (3.4%) | 0 | 56 (9.3%) | 0 | |
| ε3/ε3 | | 791 (47.7%) | 791 (84.1%) | 0 | 0 | |
| ε3/ε4 | | 544 (32.8%) | 0 | 544 (90.7%) | 0 | |
| ε4/ε4 | | 118 (7.1%) | 0 | 0 | 118 (100.0%) | |
| **Sex (Female)** | 1,658 | 908 (54.8%) | 526 (56.0%) | 329 (54.8%) | 53 (44.9%) | 0.076 |
| **Race (Cauc)** | 1,658 | 1,492 (90.0%) | 842 (89.6%) | 546 (91.0%) | 104 (88.1%) | 0.5 |
| **Age** | 1,658 | 70.82 (9.38) | 70.80 (9.70) | 71.30 (9.11) | 68.59 (7.66) | 0.001 |
| **Education (years)** | 1,658 | 15.30 (3.08) | 15.37 (3.02) | 15.20 (3.16) | 15.28 (3.17) | 0.6 |
| **Diagnosis** | 1,623 |  |  |  |  | <0.001 |
| AD |  | 476 (29.3%) | 199 (21.8%) | 230 (38.7%) | 47 (40.2%) |  |
| Control |  | 520 (32.0%) | 352 (38.6%) | 151 (25.4%) | 17 (14.5%) |  |
| MCI |  | 418 (25.8%) | 224 (24.6%) | 151 (25.4%) | 43 (36.8%) |  |
| Other dementia |  | 209 (12.9%) | 137 (15.0%) | 62 (10.4%) | 10 (8.5%) |  |
| **A status (A+)** | 980 | 556 (56.7%) | 226 (41.6%) | 265 (72.8%) | 65 (89.0%) | <0.001 |
| **T status (T+)** | 138 | 50 (36.2%) | 36 (35.0%) | 13 (39.4%) | 1 (50.0%) | 0.9 |
| **N status (N+)** | 139 | 37 (26.6%) | 28 (27.2%) | 8 (23.5%) | 1 (50.0%) | 0.6 |
| **APOE4 (NPQ)** | 1,658 | 5.19 (5.82) | 0.35 (1.65) | 11.30 (1.72) | 12.68 (2.57) | <0.001 |
| **CDR global score** | 1,591 |  |  |  |  | NA |
| 0 |  | 333 (20.9%) | 212 (23.8%) | 106 (18.2%) | 15 (12.8%) |  |
| 0.5 |  | 640 (40.2%) | 349 (39.2%) | 229 (39.2%) | 62 (53.0%) |  |
| 1 |  | 459 (28.8%) | 256 (28.8%) | 174 (29.8%) | 29 (24.8%) |  |
| 2 |  | 135 (8.5%) | 63 (7.1%) | 62 (10.6%) | 10 (8.5%) |  |
| 3 |  | 24 (1.5%) | 10 (1.1%) | 13 (2.2%) | 1 (0.9%) |  |
| **CDR Sum of Boxes** | 1,460 | 2.79 (3.45) | 2.44 (3.24) | 3.27 (3.73) | 3.24 (3.25) | <0.001 |

Continuous variables are reported as means with standard deviations (SDs), and categorical variables are summarized as counts and percentages. Total sample size (N) differed by data availability.
<sup>a</sup>P-values compare non-carriers, heterozygotes, and homozygotes using Kruska-Wallis tests for continuous variables and Pearson Chi-square or Fisher exact tests for categorical variables, as appropriate. P-values were reported as NA when sparse data or insufficient outcome variation precluded reliable statistical testing. Plasma APOE4 abundance is reported as NULISA protein quantification (NPQ). Abbreviations: A, amyloid; CDR, Clinical Dementia Rating; N, neurodegeneration; T, tau.

APOE genotypes were determined using TaqMan SNP genotyping assays targeting rs429358 and rs7412 as described.^20^ Due to strong linkage disequilibrium between these two loci, only three common APOE alleles (ε2, ε3, ε4) were present, giving rise to six possible genotype groups of ε2/ε2, ε2/ε3, ε2/ε4, ε3/ ε3, ε3/ε4, and ε4/ε4.^3^

### 2.2 Amyloid-beta pathology assessment

A subset of ADRC participants (n=739, 52.5%) and most participants for LATTICE (n=68, 88.3%), HeartSCORE (n=139, 100%) and ALPS (n=34, 100%) underwent positron emission tomography (PET) imaging for in vivo characterization of Aβ pathology using [¹¹C] Pittsburgh Compound B (PiB) as the tracer (**Supplementary Table S2**). The imaging acquisition and processing protocols have been described in detail in prior publications.^21,22^ Participants were dichotomized as amyloid positive (Aβ+) or amyloid negative (Aβ−) based on a standardized uptake value ratio (SUVR) calculated using volume weighted averaging across nine amyloid pathology-associated regions: anterior cingulate, posterior cingulate, insula, superior frontal cortex, orbitofrontal cortex, lateral temporal cortex, parietal cortex, precuneus, and ventral striatum, with SUVR >1.346 as Aβ+, <=1.346 as Aβ-.

Amyloid plaque burden in a subset of ADRC participants was assessed by postmortem neuropathological evaluation. For those participants, amyloid positivity was defined by the neuritic plaque density under the NIA-AA AD neuropathologic criteria, with CERAD scores of 2-3 considered positive and scores of 0-1 considered negative.^23^

### 2.3 NULISAseq CNS disease panel 120 assays

Plasma samples were analyzed at Department of Psychiatry, University of Pittsburgh, on an ArgoHT system, using two different versions of CNS Disease Panel 120 (Alamar Biosciences, Inc). The experimental procedures have been described in prior publications.^11–13^ In brief, thawed plasma samples were centrifuged at 10,000 × g for 10 minutes to remove particulates. Immunocomplexes were formed by incubating samples with a cocktail of paired capture and detection antibodies conjugated to target specific DNA barcodes for 127 biomarkers for version #1 (used for cohorts ALPS, HeartSCORE, and LATTICE) and 131 for version #2 (used in ADRC cohort), along with an internal control (IC). A double capture and release process was used to reduce nonspecific background binding. Following immunocomplex formation, T4 DNA ligase was used to ligate the proximal ends of DNA attached to paired antibodies and sample specific barcodes. The resulting DNA molecules were quantified by next generation sequencing (NGS). Raw NGS counts were normalized to the IC and inter plate controls, log₂ transformed, and reported as NULISA Protein Quantification (NPQ) values, which were used as a surrogate measure of protein abundance. A comprehensive list of biomarkers included in the assay, along with their corresponding protein names and gene symbols, is available in the supplementary materials (**Supplementary Table S3**).

### 2.4 Statistical analyses

All statistical analyses were performed in R version 4.4.0 (R Foundation for Statistical Computing, Vienna, Austria). Continuous variables are presented as mean and standard deviations (SDs) and categorical variables as counts and percentages. Group differences were evaluated using the Wilcoxon rank sum test or Kruskal-Wallies test for continuous variables and Pearson’s Chi-squared or Fisher’s exact test for categorial variables, as appropriate.

The discriminatory performance of plasma APOE proteotype to classify APOE ε4 genotype status was evaluated using ADRC as the discovery cohort and ALPS, HeartSCORE, and LATTICE as validation cohorts. Two classification tasks were applied: (1) APOE ε4 carrier versus non-carrier, and (2) ε4 copy number among carriers, defined as heterozygous carriers (ε2/ε4 or ε3/ε4) versus homozygous carriers (ε4/ε4), using APOE genotype as the reference standard. Receiver operating characteristic (ROC) curves were generated using plasma APOE4 level alone and with adjustment for age, sex, and race. Optimal thresholds were derived in ADRC using the Youden index and then applied without refitting to validation cohorts. Performance was summarized using area under ROC curve (AUC), sensitivity, specificity, positive predictive value (PPV), negative predictive value (NPV), accuracy, and Cohen’s κ effect size. AUC 95% CIs were estimated using 1,000 bootstrap resamples.

For association analyses, APOE ε4 status was defined either by genotype or by plasma APOE4 proteotype using the discovery-derived thresholds. Logistic regression was used to evaluate associations with Aβ positivity, defined by PiB-PET or autopsy-confirmed neuritic plaques. Models were adjusted for age, sex, and race; combined-cohort models were additionally adjusted for cohort. Analyses were performed for binary ε4 carrier status, ε4 copy number, and APOE4 proteotype quartiles among proteotype-defined carriers.

In ADRC, associations with baseline CDR Global Score (CDR-GS) were evaluated using proportional odds models adjusted for age, sex, and race. Longitudinal clinical progression was analyzed using Kaplan-Meier curves and Cox proportional hazards models. Progression was defined as any increase in CDR-SB. Cox models were adjusted for age, sex, race, and baseline CDR-SB. Hazard ratios and 95% CIs were reported for genotype- and proteotype-defined APOE ε4 carrier status, ε4 copy number, and APOE4 proteotype quartiles among proteotype-defined carriers. Likelihood ratio tests were used to compare nested Cox models. Analyses of APOE4 proteotype quartiles and ε4 homozygote subgroups were considered exploratory because of limited subgroup sizes. Statistical significance was defined as a two-sided p < 0.05.

## 3 Results

### 3.1 Study participants

A total of 1,658 participants were included from four cohorts: ADRC (n=1,408), ALPS (n=34), HeartSCORE (n=139) and LATTICE (n=77) (**Table 1, Supplementary Table S4**). The median age is 70.8 years (interquartile range [IQR]: 9.4), 54.8% were female, and 90.0% self-reported as non-Hispanic White. The APOE genotype distribution showed that 47.7% of participants were ε3/ε3, followed by ε3/ε4 (32.8%), ε2/ε3 (8.5%), ε4/ε4 (7.1%), ε2/ε4 (3.4%), and ε2/ε2 (0.5%). Overall, 43.3% of participants carried at least 1 APOE ε4 allele.

As expected, APOE ε4 carriers were most frequent in the ADRC memory clinic cohort (45.8%), which includes a large proportion of individuals with cognitive impairment/dementia, followed by LATTICE (39.0%), a lithium treatment trial targeting individuals with mild cognitive impairment, and then the community-based HeartSCORE (25.9%) and ALPS (20.6%) cohorts, which consist predominantly of cognitively normal individuals. Among participants with available amyloid-beta (Aβ) pathology (PiB-PET or autopsy), 56.7% were Aβ+, and being ε4 carriers was associated with significantly higher Aβ+ rate than non-carriers (75.5% vs. 41.6%; p < 0.001). Among the carriers, homozygous carriers were associated with significantly higher Aβ+ rate (89.0% in homozygous vs. 72.8 in heterozygous; p = 0.003), suggesting dose-dependent penetrance of ε4 allele on Aβ pathology. In addition, APOE ε4 carriers were also associated with increased risk of having mild cognitive impairment (MCI) or AD (p < 0.001), also worsen in homozygous than heterozygous carriers (p = 0.019).

Boxplots of APOE4 proteotype distribution by carriership and allele dosage are shown in **Supplementary Fig. S1**. The majority of genotyped non-carriers exhibited low APOE4 NPQ values (<5), whereas most carriers had NPQ values >5, with a small number of ADRC samples (14 out of 645 carriers; 18 out of 763 non-carriers) deviating from this pattern. A stepwise increase in APOE4 NPQ values was observed across non-carriers, heterozygotes, and homozygotes (Kruskal-Wallis H(2) = 1315.6, p < 0.001). When cohorts were compared among APOE non-ε4 carriers, NPQ values differed significantly across cohorts (H(3) = 23.59, p < 0.001). This difference remained significant after excluding 18 ADRC participants with relatively high NPQ values (H(3)=34.34, p < 0.001), likely reflecting assay batch effect. Among APOE ε4 heterozygotes, NPQ values were slightly higher overall in LATTICE, but differences across cohorts were not statistically significant (within heterozygotes p = 0.083).

### 3.2 Plasma APOE4 proteotype accurately predicts APOE ε4 carriership

We first tested whether plasma APOE4 proteotype could classify APOE ε4 carrier status. The ADRC cohort was used for threshold discovery because of its large sample size and broad range of cognitive severity; ALPS, HeartSCORE, and LATTICE were used as independent validation cohorts. **Fig. 1A-B** shows the ROCs for discriminating APOE ε4 carriers vs. non-carriers and heterozygous carriers vs. homozygous carriers in the discovery cohort, respectively. Contingency tables and classification performance for the three validation cohorts are shown in **Fig. 1C-D**, with detailed classification metrics listed in **Supplementary Table S5**.

**Figure 1.**
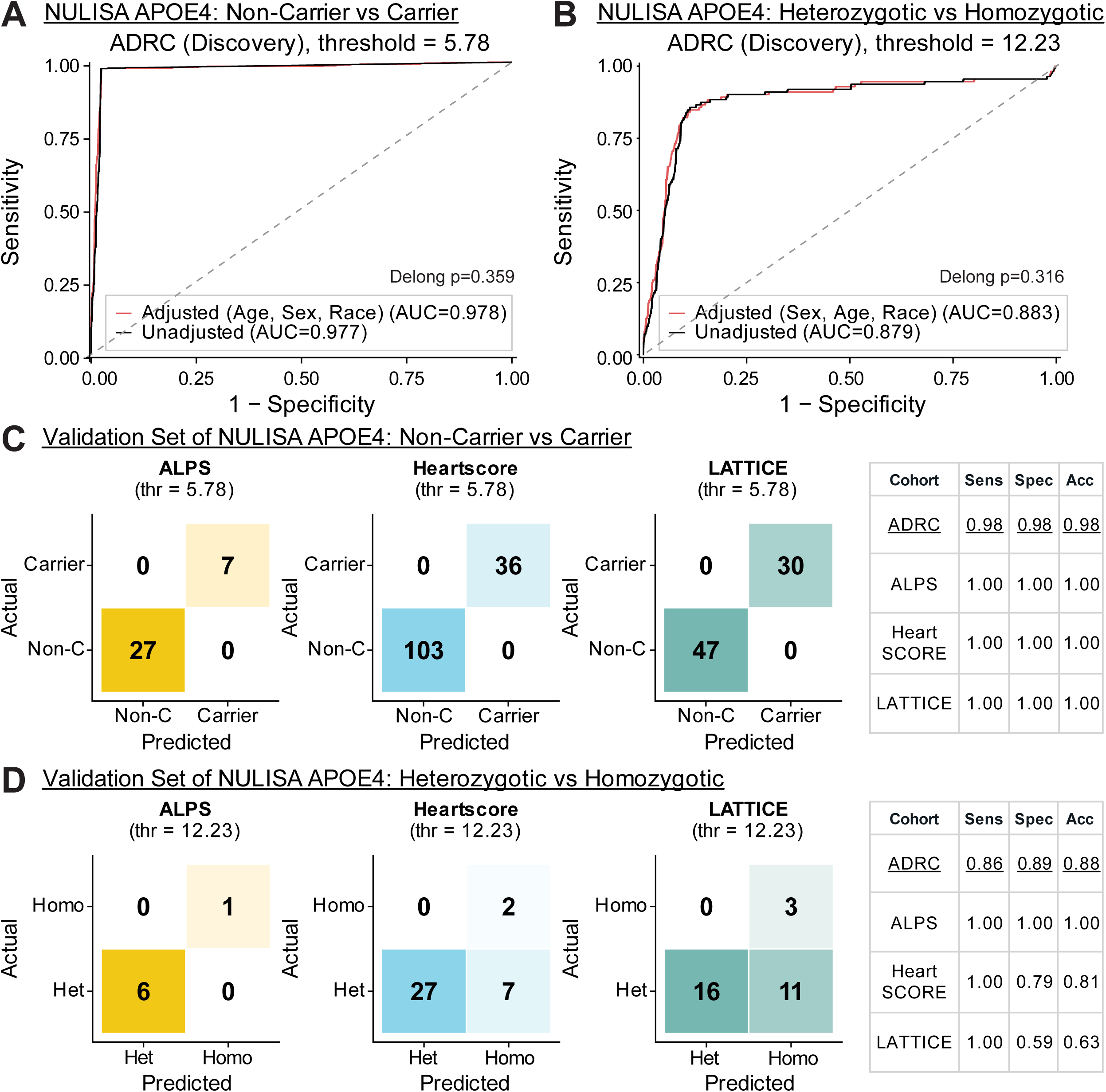
Concordance between NULISA APOE4 proteotype and APOE genotype. (**A-B**) Receiver operating characteristic curves show the ability of NULISA APOE4 proteotype in discriminating APOE ε4 carriership (**A**) and allele dosage (**B**) in the ADRC discovery cohort, with (red curves) and without the inclusion of covariates (age, sex, and race; black curves). DeLong tests compared adjusted and unadjusted AUCs. (**C-D**) Contingency tables and performance metrics in the validation cohorts for differentiating carriers vs. non-carriers (**C**) and heterozygous carriers vs. homozygous carriers (**D**). Abbreviations: thr, Youden-index-derived thresholds; Sens, sensitivity; Spec, specificity; Acc, accuracy; Het, heterozygous carriers; Homo, homozygous carriers; Non-C, non-carrier.

Plasma APOE4 proteotype predicted both APOE ε4 carriership and allele dosage with high accuracy (**Fig. 1A**). In the ADRC discovery cohort, plasma APOE4 proteotype near-perfect discrimination between APOE ε4 carriers and non-carriers, with an area under the curve (AUC) of 0.977 when used alone and 0.978 after adjustment of age, sex and race (**Fig. 1A**). A Youden-index-based NPQ threshold of 5.78 yielded 97.8% sensitivity and 97.6% specificity. Inclusion of covariates, age, sex, and race did not statistically improve the performance (DeLong p=0.359). Notably, applying this same threshold to the three independent validation cohorts resulted in 100% accuracy in distinguishing APOE ε4 carriers from non carriers (**Fig. 1C**).

In differentiating APOE ε4 homozygotes from heterozygotes, plasma APOE4 prototype alone achieved an AUC of 0.879 in the ADRC discovery cohort. Inclusion of covariates resulted in only slight and non-significant increase in AUC. Classification using Youden index determined NPQ threshold of 12.23 led to 85.7% sensitivity and 88.7% specificity (**Fig. 1B and Supplementary Table S5**). Applying the same threshold to the validation cohorts resulted in accuracies of 100%, 80.6% and 63.3%, in ALPS, HeartSCORE, and LATTICE, respectively (**Fig. 1D**). The main source of errors fell among heterozygous APOE ε4 carriers being classified as homozygous by plasma APOE4 proteotype. We examined whether these discordant classifications were associated with clinical differences. As shown in **Supplementary Fig. S2**, no significant differences were observed in clinical measures, including Clinical Dementia Rating (CDR) Global Score (CDR-GS), clinical AD diagnosis, or neuroimaging-based ATN classification.

It is worth noting that, although a Youden index–derived threshold of 5.78 NPQ value was used for performance evaluation, there was no overlap in NPQ values between APOE ε4 carriers and non-carriers for all three validation cohorts. Any threshold within the range of 2.78 to 8.43 would yield identical classification performance.

Several ADRC samples (n=18) classified as non-carriers by genotype exhibited high APOE4 NPQ values. To assess whether these proteotype–genotype discordant cases influenced performance, we repeated the discovery analysis after excluding these discordant individuals. The optimal thresholds remain unchanged for both carriership and allele dosage classification (non-carriers vs carriers: 5.78 NPQ value; heterozygous vs homozygous: 12.23 NPQ value). AUCs also remain stable, with overlapping 95% CIs (full vs filtered ADRC: 0.977 vs 1.000 [Non-C/Carrier] and 0.880 vs 0.910 [Het/Homo]; **Supplementary Fig. S3**). These results indicate that discordant cases did not drive the observed discrimination.

### 3.3 Plasma APOE4 proteotype shows genotype-like associations with Aβ positivity

We next evaluated whether the plasma APOE4 proteotype showed comparable associations with Aβ positivity relative to APOE genotype. These analyses were restricted to cohorts with at least 10 APOE ε4 carriers for carrier-status analyses and at least 5 homozygous carriers for copy number-based analyses, with detailed number of subjects included in each analysis listed in **Supplementary Table S6**. Consistent with the high agreement between proteotype- and genotype-defined APOE ε4 carriership, the two approaches showed nearly identical associations with Aβ positivity (**Fig. 2A**).

**Figure 2.**
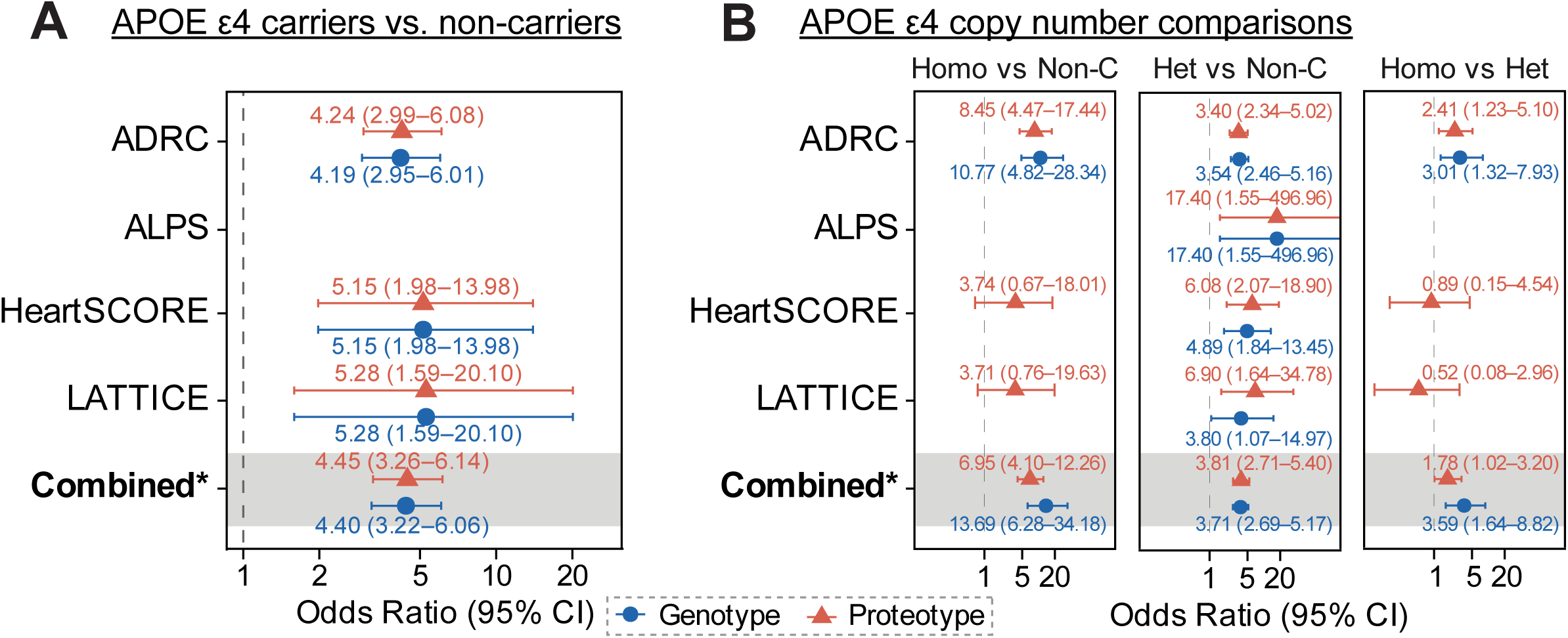
Comparison of APOE4-associated risk for Aβ positivity by genotype and proteotype. Forest plots show odds ratios and 95% confidence intervals for Aβ positivity in the ADRC, ALPS, HeartSCORE, LATTICE, and combined cohorts. Within each panel, cohort names correspond to the estimates displayed on the same horizontal row. (**A**) Odds ratios compared APOE ε4 carriers with non-carriers. (**B**) Odds ratios compared three copy-number comparisons, from left to right: homozygous carriers versus non-carriers, heterozygous carriers versus non-carriers, and homozygous versus heterozygous carriers. Models were adjusted for sex, age, and race; combined analyses additionally included cohort as a covariate. Blue circles indicate APOE ε4 genotype groups, and red triangles indicate groups defined by plasma APOE4 proteotype using Youden-derived thresholds (5.78 NPQ values for distinguishing carriers from non-carriers, and 12.23 NPQ values for distinguishing heterozygous from homozygous carriers. Horizontal bars denote 95% confidence intervals vertical dashed line indicates an odds ratio of 1 (no risk difference). Abbreviations: APOE, apolipoprotein E4; Aβ, amyloid-PET; NULISA, nucleic acid-linked immuno-sandwich assay; ADRC, Alzheimer’s Disease Research Center; ALPS, Alzheimer’s Pathways Sleep Study; HeartSCORE, Heart Strategies Concentrating on Risk Evaluation; LATTICE, Lithium as a Treatment to Prevent Impairment of Cognition in Elders; NPQ, NULISA protein quantification; CI, confidence interval; Het, heterozygous carriers; Homo, homozygous carriers; Non-C, non-carriers.

In ADRC, among 739 participants with complete covariate and Aβ data, genotype-defined APOE ε4 carriers were associated with higher odds of Aβ positivity (odds ratio [OR]=4.19; 95% CI: 2.95-6.01), after adjustment for age, sex, and race. A nearly identical OR was observed for APOE4 proteotype-defined carrier status (OR=4.24; 95% CI: 2.99-6.08). Similar concordance between genotype- and proteotype-based estimates was observed in the other eligible cohorts and in the combined sample after adjustment for cohort (**Fig. 2A**).

We then compared genotype- and proteotype-based APOE ε4 dosage with Aβ positivity. As shown in **Fig. 2B**, in ADRC, genotype-defined heterozygous APOE ε4 carriers were associated with an OR of 3.54 (95% CI: 2.46-5.16), while homozygous APOE ε4 carriers had an OR of 10.77 (95% CI: 4.82-28.34) and 3.01 (95% CI: 1.32-7.93) compared to non-carriers and heterozygotes, respectively. Correspondingly, the ORs based on proteotype-defined status were 3.40 (95% CI: 2.34-5.02), 3.54 (95% CI: 2.46-5.16), and 8.45 (95% CI: 4.47-17.44), respectively, and were of a similar magnitude to those observed using genotype-based classification.

### 3.4 APOE4 proteotype mirrors APOE genotype associations with clinical manifestations

We next evaluated whether APOE ε4 status defined by genotype or proteotype showed similar associations with cognitive status and longitudinal clinical progression in ADRC, the cohort with the most complete clinical information. As shown in **Fig. 3**, APOE ε4 carriers defined by either genotype or proteotype had higher CDR-GS, indicating worse cognitive status. Both measures showed similar effects, with a proportional odds ratio of 1.5, indicating a consistent increase in odds of greater cognitive impairment across CDR-GS levels. Similar findings were observed when stratifying participants into non-carriers, heterozygous carriers, and homozygous carriers. The proportional odds ratios for heterozygous versus non-carriers were 1.49 and 1.47 for genotype- and proteotype-defined APOE ε4 status, respectively, whereas those for homozygous versus non-carriers were 1.48 and 1.60.

**Figure 3.**
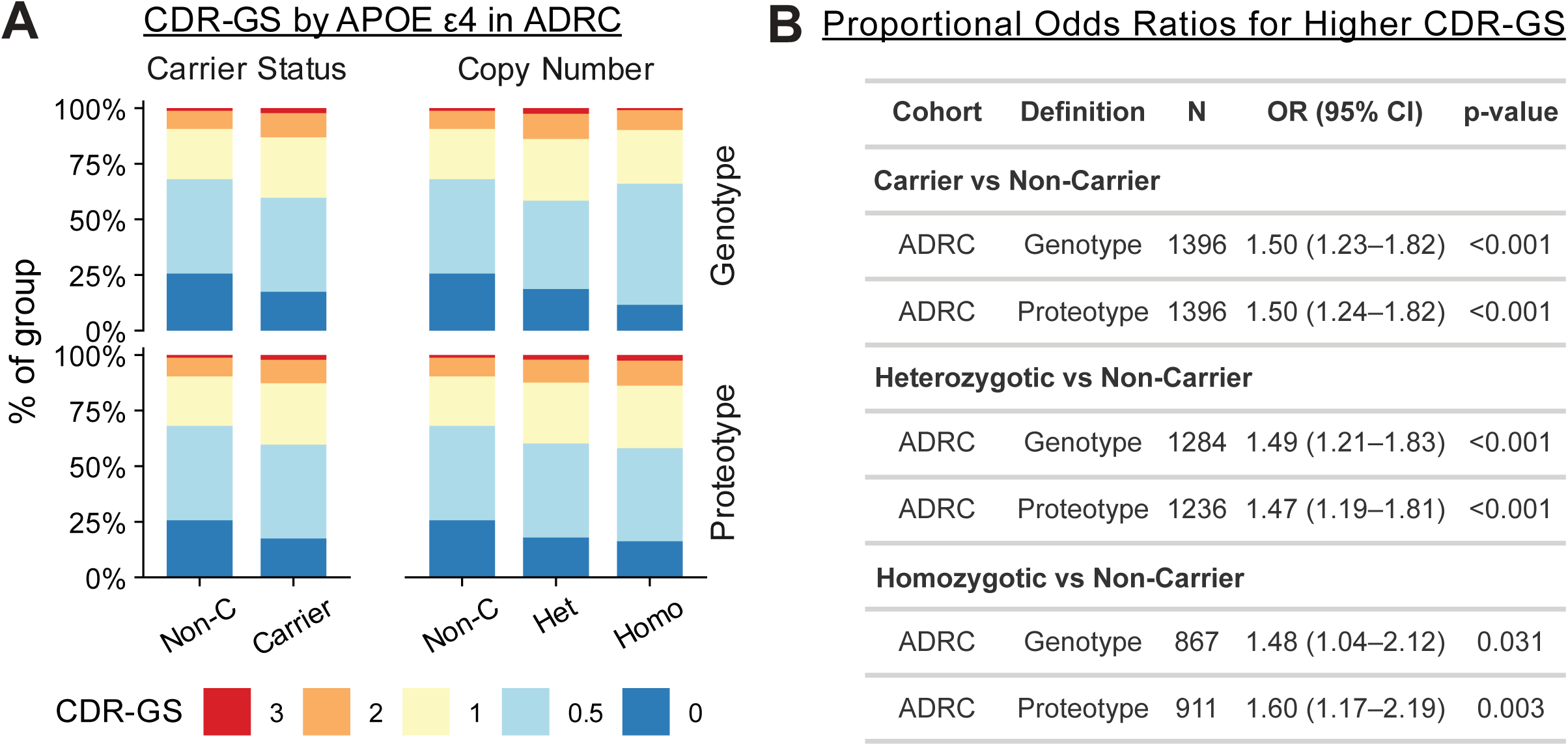
APOE ε4 genotype and proteotype associations with CDR-GS in ADRC. **(A**) Distribution of CDR-GS categories by APOE ε4 carrier status and APOE ε4 copy number in the ADRC cohort, shown separately for genotype-defined and proteotype-defined APOE ε4 status. Stacked bars indicate the percentage of participants in each CDR-GS category. (**B**) Proportional odds ratios for higher CDR-GS according to APOE ε4 carrier status and copy number. Models compare carriers, heterozygotes, and homozygotes with non-carriers and are adjusted for age, sex, and race. Abbreviations: CDR-GS: clinical dementia scale-global score; ADRC, Alzheimer’s Disease Research Center; OR, odds ratio; CI, confidence interval; Het, heterozygous carriers; Homo, homozygous carriers; Non-C, non-carriers.

When examining the association of APOE ε4 status with CDR sum of box (CDR-SB)-defined cognitive trajectories, APOE ε4 carriers defined by genotype and proteotype also showed nearly identical accelerated decline rate over non-carriers, with hazards ratio (HR) of 1.22 (95%CI: 1.06-1.41) and 1.22 (95%CI: 1.05-1.40), respectively (**Table 2A**, **Fig. 4 A-B**). Likelihood ratio test indicated no significant difference between the two HRs, with p = 0.971.

**Table 2.**
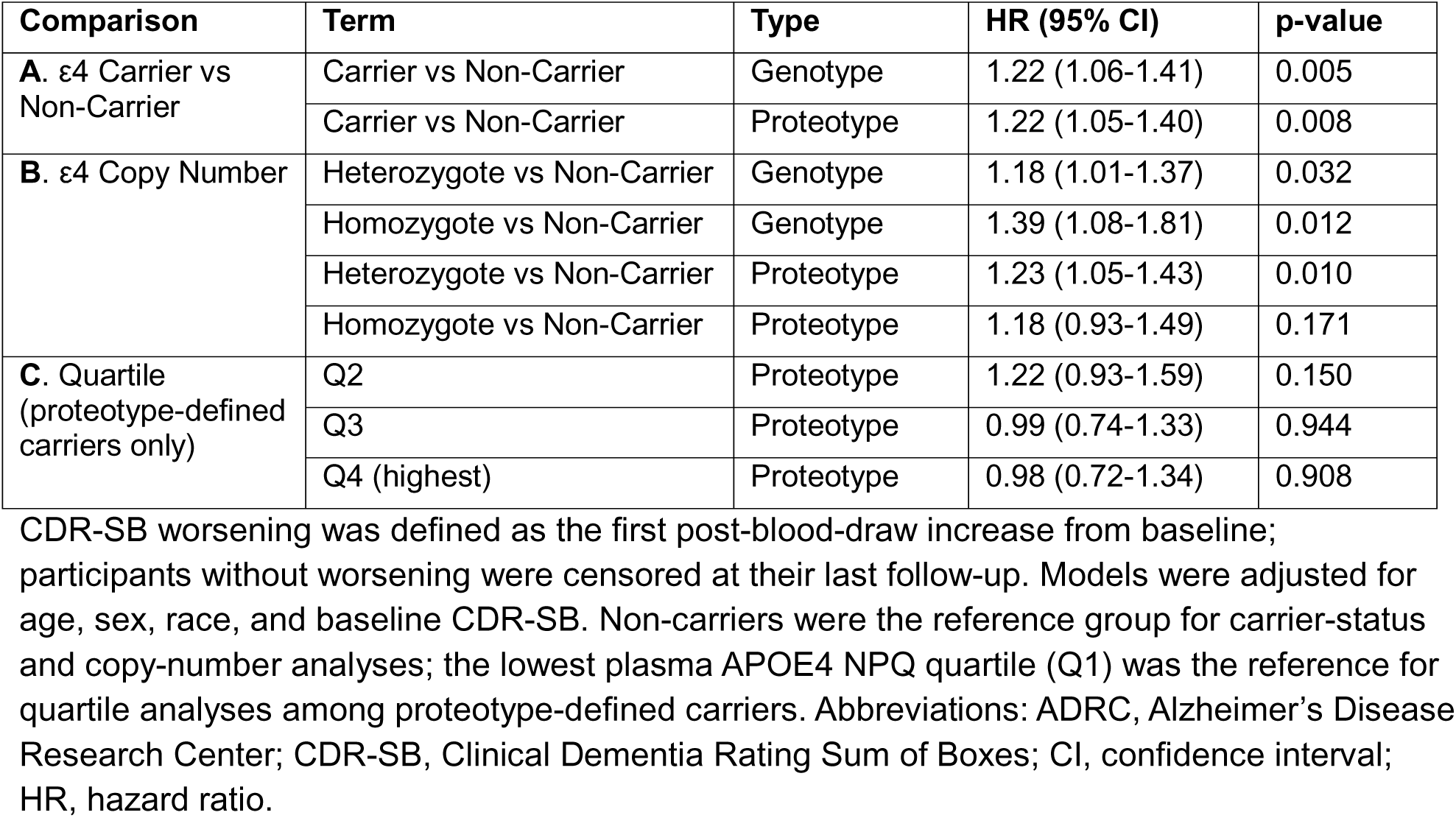
Associations of APOE ε4 genotype- and proteotype-defined groups with time to CDR-SB worsening in ADRC.

| Comparison | Term | Type | HR (95% CI) | p-value |
| --- | --- | --- | --- | --- |
| **A. ε4 Carrier vs Non-Carrier** | Carrier vs Non-Carrier | Genotype | 1.22 (1.06-1.41) | 0.005 |
|  | Carrier vs Non-Carrier | Proteotype | 1.22 (1.05-1.40) | 0.008 |
| **B. ε4 Copy Number** | Heterozygote vs Non-Carrier | Genotype | 1.18 (1.01-1.37) | 0.032 |
|  | Homozygote vs Non-Carrier | Genotype | 1.39 (1.08-1.81) | 0.012 |
|  | Heterozygote vs Non-Carrier | Proteotype | 1.23 (1.05-1.43) | 0.010 |
|  | Homozygote vs Non-Carrier | Proteotype | 1.18 (0.93-1.49) | 0.171 |
| **C. Quartile (proteotype-defined carriers only)** | Q2 | Proteotype | 1.22 (0.93-1.59) | 0.150 |
|  | Q3 | Proteotype | 0.99 (0.74-1.33) | 0.944 |
|  | Q4 (highest) | Proteotype | 0.98 (0.72-1.34) | 0.908 |

CDR-SB worsening was defined as the first post-blood-draw increase from baseline; participants without worsening were censored at their last follow-up. Models were adjusted for age, sex, race, and baseline CDR-SB. Non-carriers were the reference group for carrier-status and copy-number analyses; the lowest plasma APOE4 NPQ quartile (Q1) was the reference for quartile analyses among proteotype-defined carriers. Abbreviations: ADRC, Alzheimer's Disease Research Center; CDR-SB, Clinical Dementia Rating Sum of Boxes; CI, confidence interval; HR, hazard ratio.

**Figure 4.**
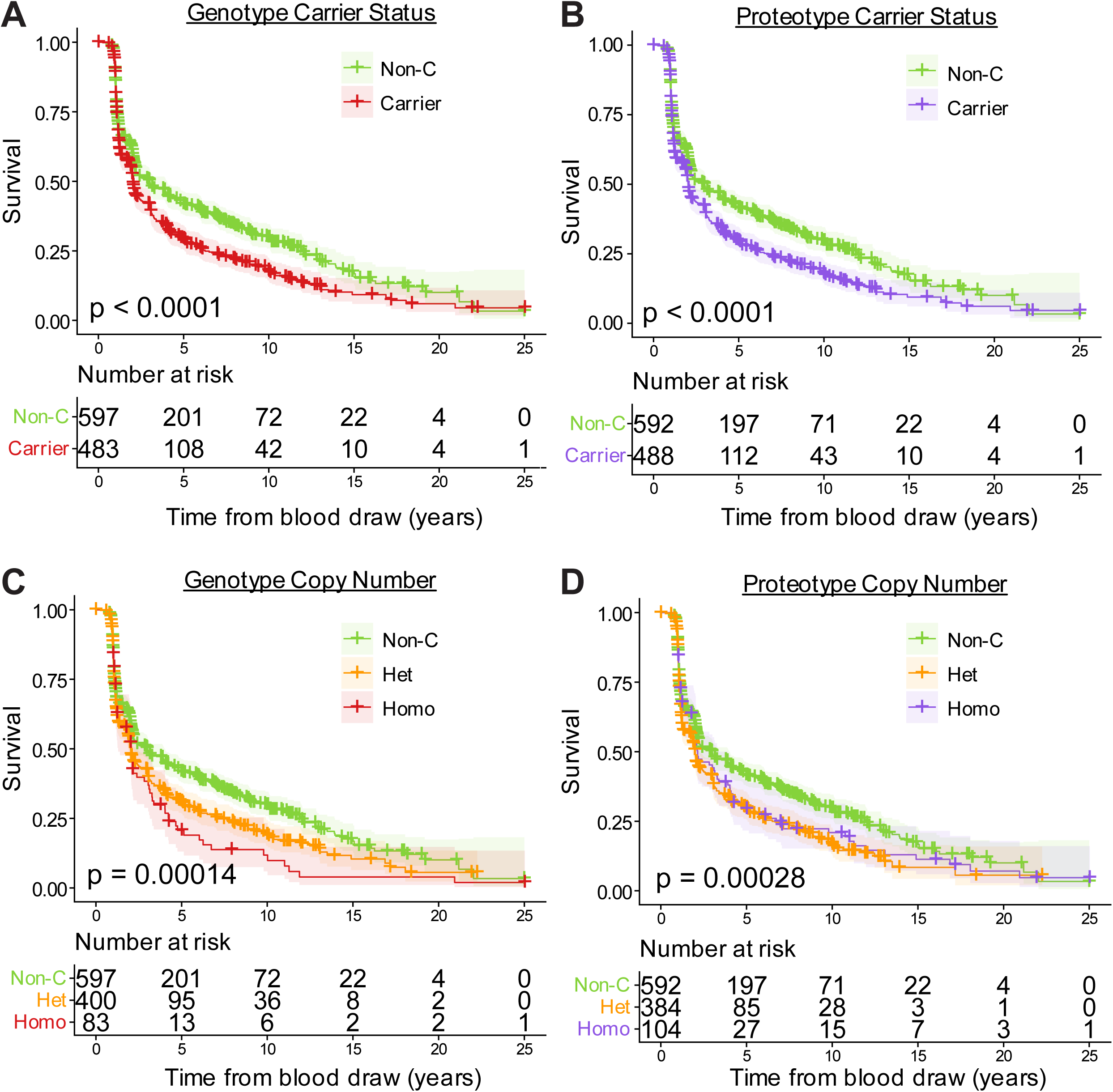
Kaplan-Meier curves of APOE ε4 genotype and proteotype with CDR-SB worsening in ADRC. (**A-B**) Survival curves for time free of CDR-SB worsening by genotype-defined and proteotype-defined APOE ε4 carrier status. (**C-D**) Survival curves for time free of CDR-SB worsening by genotype-defined and proteotype-defined APOE ε4 copy number. P values are obtained from log-rank group comparisons of survival distributions. Abbreviations: CDR-SB, Clinical Dementia Rating Sum of Boxes; Het, heterozygous carriers; Homo, homozygous carriers; Non-C, non-carriers.

Copy-number analyses showed a clearer dose-response pattern for genotype-defined APOE ε4 groups. Compared with non-carriers, genotype-defined heterozygotes had an HR of 1.18 (95% CI, 1.01-1.37), and genotype-defined homozygotes had an HR of 1.39 (95% CI, 1.08-1.81) (**Fig. 4C**; **Table 2A**). For proteotype-defined groups, heterozygotes had increased risk of CDR-SB worsening compared with non-carriers (HR, 1.23; 95% CI, 1.05-1.43), whereas the association for proteotype-defined homozygotes was not statistically significant (HR, 1.18; 95% CI, 0.93-1.49) (**Fig. 4D**; **Table 2B**).

## 4 Discussion

In this multi-cohort study, plasma NULISA APOE4 closely recapitulated the clinical value of APOE ε4 genotype. APOE4 proteotyping classified ε4 carrier status with near-perfect accuracy in the ADRC discovery cohort and retained perfect external validation for carrier classification across ALPS, HeartSCORE, and LATTICE, supporting its validity as a blood-based surrogate of traditional APOE genotyping. APOE proteotype also performed comparably to conventional genotyping as a standalone predictor of amyloid positivity and cognitive impairment in both cross-sectional and longitudinal analyses.

Several APOE proteotyping assays, including both mass spectrometry-based^24,25^ and immunoassay-based^26–28^ approaches, have been developed, and their clinical feasibility as surrogates for APOE genotyping has been demonstrated in multiple prior studies. Our results, derived from multiple cohorts spanning preclinical, community-based populations to memory clinic settings, further reaffirm the clinical utility of APOE proteotyping. The APOE proteotyping assay evaluated in this study is an ultrasensitive immunoassay integrated into a multiplex platform. Dittrich et al. previously assessed the use of pan APOE and APOE4, both included in the NULISA CNS panel, for APOE proteotyping.^14^ Since then, the NULISA CNS panel has undergone updates, with the pan apoE assay now exhibiting hook effect–related behavior, which complicates the use of the APOE4-to–pan APOE ratio for proteotyping. Our findings demonstrate that APOE4 alone can achieve similarly high accuracy as a surrogate for APOE genotyping.

These findings have a practical clinical implication. APOE-related risk information can be obtained directly from a plasma proteomic workflow rather than requiring a separate DNA-based test, and that same blood draw can simultaneously provide additional disease-relevant biomarker information that is more informative for amyloid risk assessment than APOE genotype alone. From a translational perspective, this shifts APOE from a standalone genetic result to an integrated component of a multiplex blood-based risk model. The data therefore support plasma APOE4 proteotyping as a clinically useful surrogate for APOE genotyping and support multiplex NULISA proteotyping as a more informative approach to amyloid prediction than APOE genotyping alone.

The pattern of results also helps clarify the context in which plasma APOE4 may add value. Beyond its ability to infer APOE genotype with high accuracy, APOE proteotype showed risk-prediction performance comparable to genotype when incorporated into models of amyloid positivity and cognitive worsening. These findings suggest that plasma APOE4 captures a substantial component of APOE-associated disease risk and may function as a blood-based proxy for genetic information within broader multiplex plasma biomarker panels. However, interpretation should remain cautious, as the biological consequences of APOE genotype are mediated through the encoded protein, and protein-level measures may contribute risk-relevant information that was not fully resolved in the present analyses.^29^

We identified a small subset of samples with discordant APOE genotype and proteotype, all from the ADRC cohort. This discrepancy may stem from true biological heterogeneity, such as cell-type-specific shifts in APOE expression, isoform-specific degradation rates, or post-translational modifications that mask proteotypic peptides during mass spectrometry detection.^30–32^ If driven by biology, these discrepancies imply that a patient’s genetic risk profile may not always mirrors their functional, circulating protein architecture. However, pre-analytical factors such as sample mishandling cannot be excluded. Given that pre-analytical variability has been estimated to account for over 60% of total analytical error^33^, these findings underscore the importance of minimizing handling steps to reduce assay-related variability. In this context, APOE proteotyping—particularly when implemented within an integrated, high-throughput platform—offers a practical advantage by reducing the need for additional sample processing and external genotyping workflows, thereby potentially mitigating sources of pre-analytical error.

This study has several strengths. First, the analysis was performed across multiple independent cohorts and included two assay versions, increasing confidence that the observed genotype-biomarker relationships are reproducible rather than cohort- or assay-specific. Second, the use of a discovery-testing framework with thresholds selected in ADRC and then applied to external cohorts provides a more rigorous assessment of generalizability than cohort-specific optimization. Third, we extended the comparison between genotype and proteotype to their associations with clinical AD manifestations, including amyloid positivity, cognitive impairment, and cognitive trajectories.

Several limitations should also be considered. Sample sizes for APOE ε4 homozygotes were small in external cohorts, particularly ALPS and HeartSCORE, limiting the precision and stability of heterozygous-versus-homozygous classification estimates. This sample constraint is a direct consequence of population genetics that the APOE ε4 frequency sits around 14% in European populations and mathematically yielding an expected homozygous carrier prevalence of only 2% under Hardy-Weinberg equilibrium in general populations.^3^ Accordingly, the perfect or near-perfect performance observed in some small validation subsets should be interpreted cautiously. In addition, the current assay is limited to detecting the APOE4 proteoform and therefore cannot capture information on the ε2 allele. This limits its ability to distinguish among non-carriers (ε2/ε2, ε2/ε3, ε3/ε3) and to differentiate ε4 heterozygotes (ε2/ε4 vs ε3/ε4). Finally, although cohort adjustment was used in regression models, residual differences in participant characteristics, sample collection, and assay conditions across studies may still influence measured levels.

In summary, our findings support APOE proteotyping as a surrogate for APOE ε4 genotype, with performance comparable to genotyping in predicting key Alzheimer’s disease–related outcomes across multiple cohorts. While these findings support its feasibility as a blood-based surrogate, limitations such as reduced resolution for non ε4 alleles and cohort-specific variability should be considered. Nevertheless, the ability to integrate APOE status into multiplex biomarker workflows offers a compelling advantage, particularly for large-scale and resource-limited settings. Further validation in diverse populations and clinical contexts will be important to fully establish its role alongside conventional genotyping.

## Data Availability

All data produced in the present work are contained in the manuscript

## Acknowledgements

The authors are thankful to the Pitt-ADRC, ALPS, HeartSCORE and LATTICE participants and study coordinators whose dedication and commitment were fundamental to the successful implementation of this study.

## Competing Interests Statement

XZ and TKK are inventors on University of Pittsburgh patents and provisional patents regarding biofluid biomarker methods, targets and reagents/compositions, that may generate income for the institution and/or self should they be licensed and/or transferred to other organization(s). TKK has served as an adhoc consultant and/or advisory board member for Quanterix Corporation, SpearBio Inc., Neurogen Biomarking LLC., Alzheon, Siemens Healthineers and Neurogen Biomarking LLC., outside the submitted work. TKK has received royalties from Bioventix for the transfer of specific antibodies and blood biomarker assays to third party organizations. The other authors have nothing to declare.

## Author Contributions Statement

Conception, experimental design, and supervision: XZ, TKK, RS; Cohort recruitment and retention, clinical assessments, coordination and data acquisition: OLL, ADC, JEE, AGG, KAW, SER, AS; Plasma sample preparation, biobanking and biomarker data acquisition and processing: XZ, LTC, MFF, JEE; Data compilation, statistical analysis and interpretation of data: RS, XZ. All authors contributed to manuscript draft and/or revision.

## Funding Sources

National Institutes of Health (R01AG068001, R01AG055389, R01AG068001, R01AG055389, RF1AG052525); the Anbridge Charitable Fund; Department of Psychiatry, University of Pittsburgh.

## Figure legends

**Supplementary Fig. S1. Plasma APOE4 protein levels by APOE ε4 genotype** Boxplots show NULISA-measured plasma APOE4 levels, reported as NPQ, across APOE ε4 genotype groups in ADRC, ALPS, HeartSCORE, LATTICE, and the combined cohorts. Individual points represent participants. In the upper row, participants are classified as non-carriers and carriers. In the lower row, participants are classified as non-carriers, heterozygous carriers, and homozygous carriers. Dashed horizontal lines indicate youden-index-based APOE4 NPQ reference thresholds used to distinguish genotype groups, with the green dashed line denotes the lower threshold of 5.78 NPQ for distinguishing non-carriers from carriers, and the purple dashed line denotes the upper threshold of 12.23 NPQ for distinguishing homozygotes carriers from heterozygotes carriers. Point colors indicate NULISA-defined APOE4 protein groups based on NPQ thresholds: green, below the lower threshold; gray, between the lower and upper thresholds; and purple, above the upper threshold. Abbreviations: APOE, apolipoprotein E4; NULISA, nucleic acid-linked immuno-sandwich assay; NPQ, NULISA protein quantification; ADRC, Alzheimer’s Disease Research Center; ALPS, Alzheimer’s Pathways Sleep Study; HeartSCORE, Heart Strategies Concentrating on Risk Evaluation; LATTICE, Lithium as a Treatment to Prevent Impairment of Cognition in Elders.

**Supplementary Fig. S2. Clinical distributions among genotype-proteotype concordant and discordant APOE ε4 heterozygous carriers defined by genotype** Clinical characteristics are compared between genotype-defined APOE ε4 heterozygous carriers within genotype-proteotype concordant classification and discordant classification. Panels show distribution of CDR global score (**A**), clinical diagnosis (**B**), and ATN biomarker positivity (**C**).

**Supplementary Fig. S3. ROC-based classification performance after excluding discordant APOE ε4 non-carriers.** Sensitivity analysis evaluating the impact of proteotype-genotype discordant APOE ε4 non-carriers, defined by genotype, in the ADRC cohort. Receiver operating characteristic (ROC) curves are shown for discriminating APOE ε4 carriers from non-carriers and heterozygous from homozygous carriers, with and without excluding discordant non-carriers.

## Notes

### Competing Interest Statement

The authors have declared no competing interest.

### Author Declarations

Data were drawn from the ADRC, HeartSCORE, LATTICE and ALPS studies. All four studies were approved by the University of Pittsburgh Institutional Review Board, and all participants provided written informed consent.

