## Supplementary figures and images for "Clinical utility of plasma APOE proteotyping for Alzheimer’s disease risk and prognosis"

### Supplementary Figure 1

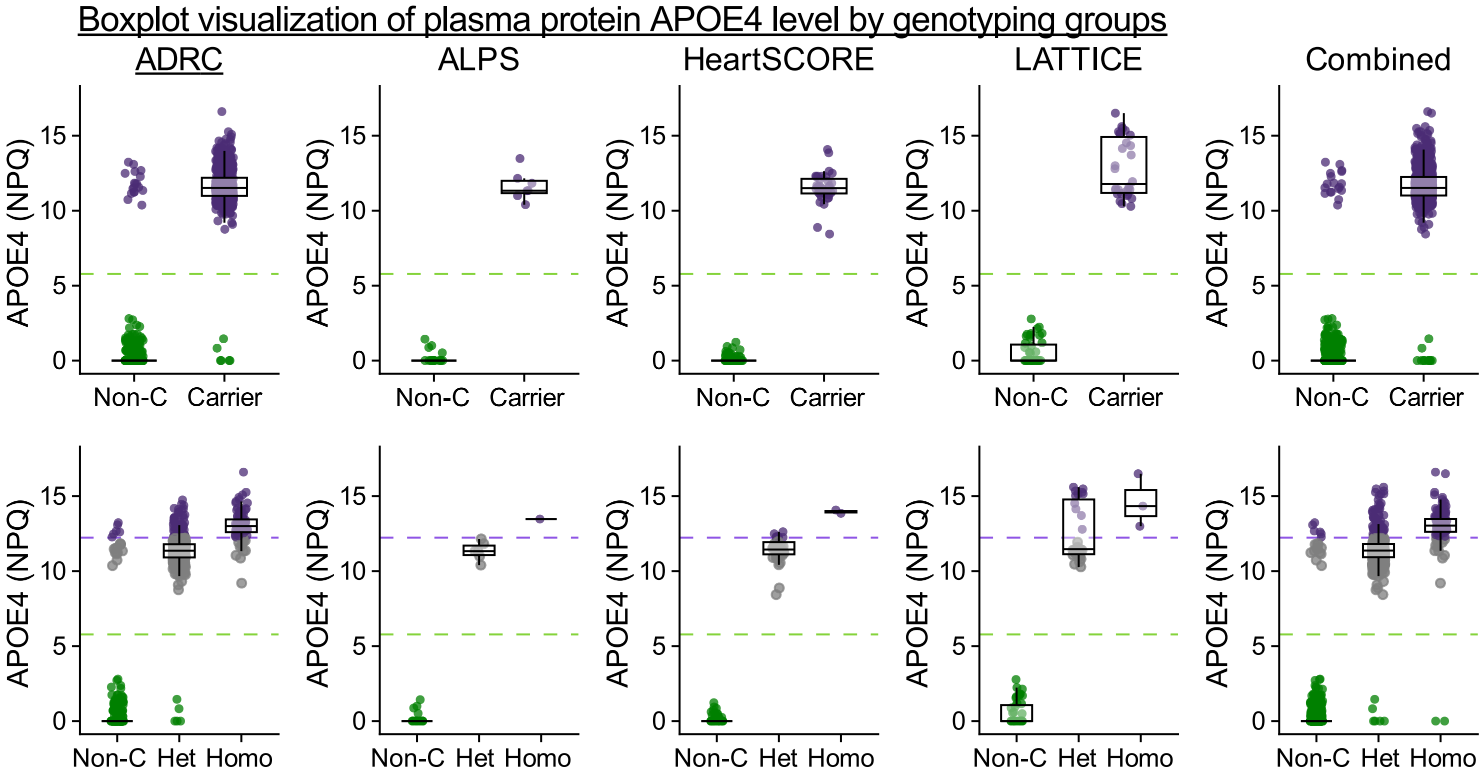

### Supplementary Figure 2

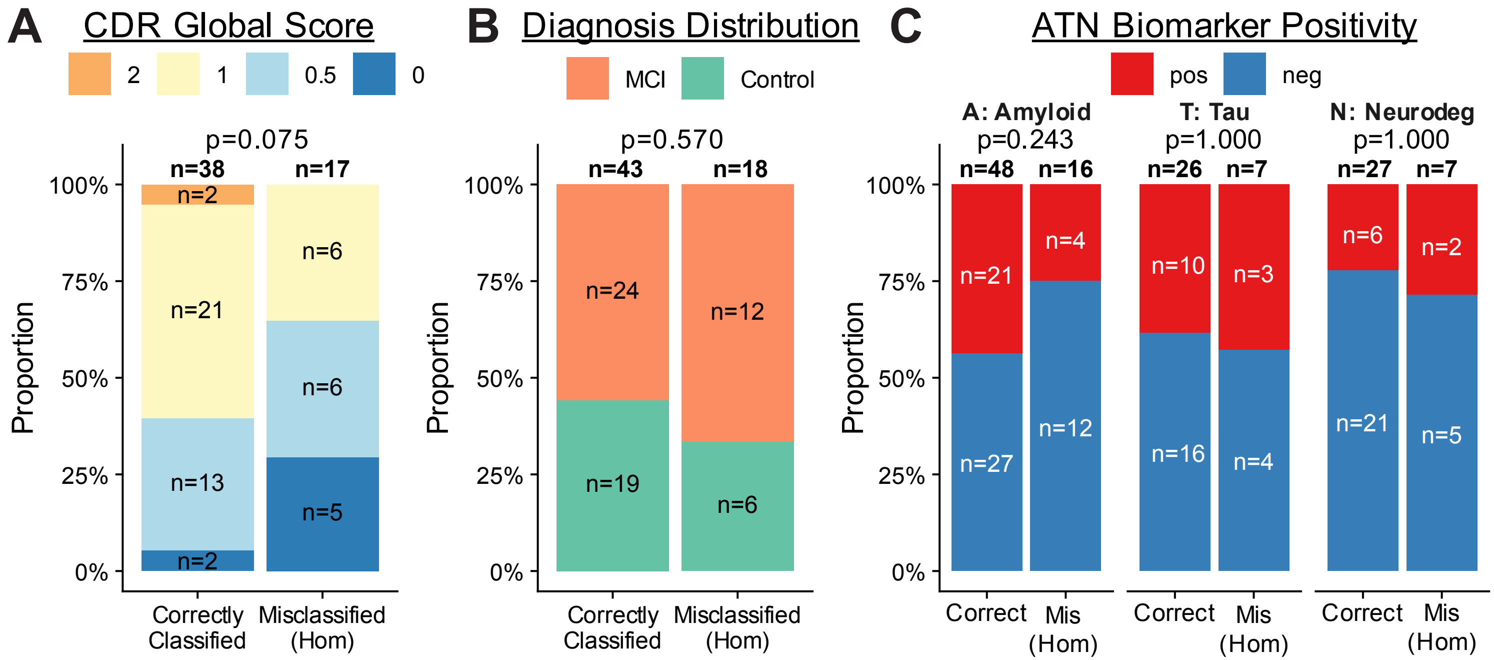

### Supplementary Figure 3

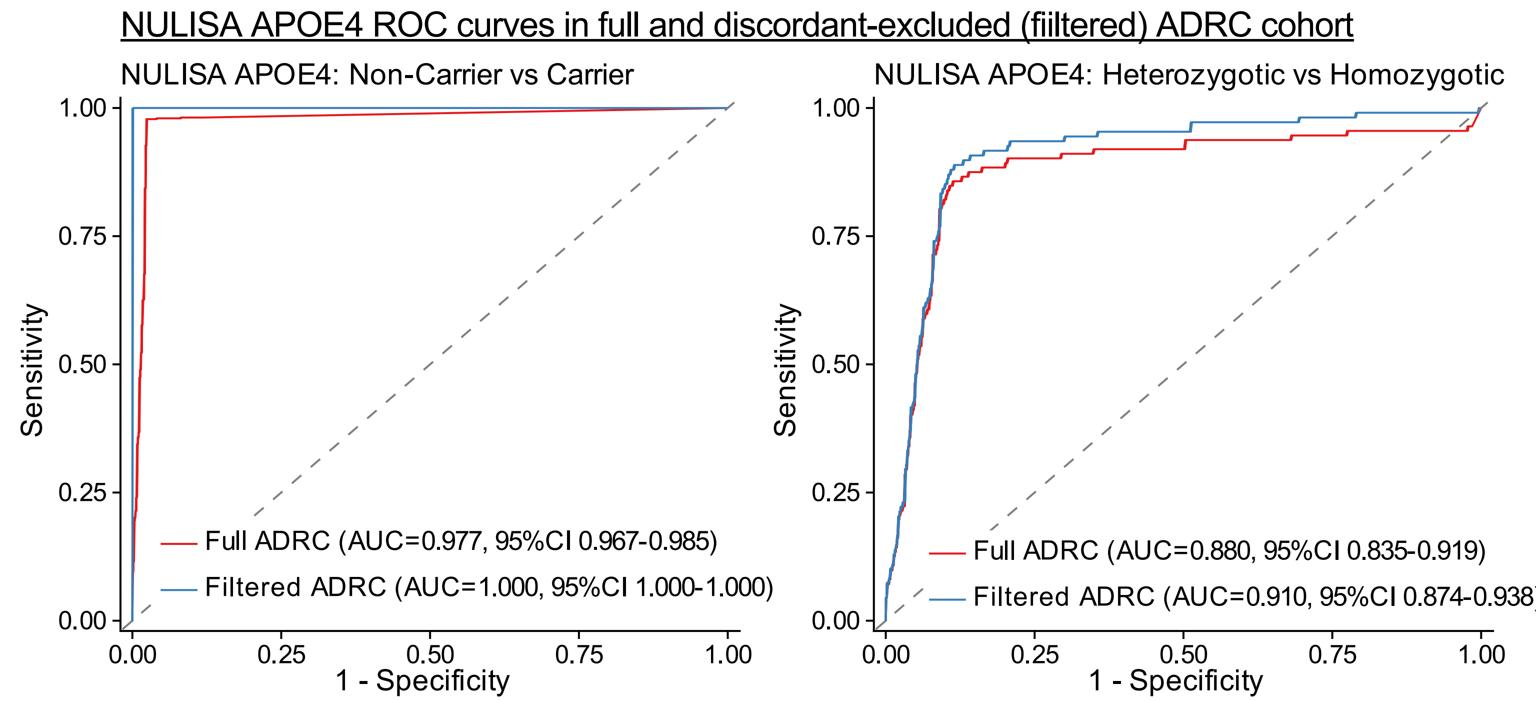
